# Signals across the pond: Bilateral airplane wastewater monitoring paves the way for international cooperation on pathogen surveillance for public health

**DOI:** 10.64898/2026.02.09.26345757

**Authors:** Matthew J. Wade, Ian Ruskey, Elizabeth Perry, Valerie Meehan, Andrew P. Rothstein, Dawn Gratalo, Sarah Rush, Birgitte Simen, UKHSA Laboratory Team, Cindy R. Friedman

## Abstract

We present findings from the first known pilot study of transatlantic airplane wastewater monitoring, conducted over six months at two connected international airports in the United States and the United Kingdom. This study demonstrates the feasibility of implementing bilateral wastewater-based pathogen surveillance at international travel hubs. We outline the operational and analytical methodologies employed, highlight key challenges encountered in transnational coordination, and provide recommendations for the design and implementation of future surveillance programs at points of entry.

## Introduction

Aviation wastewater monitoring offers the potential to enhance cross-border biosecurity. International air travel can rapidly disseminate pathogens^1^ making surveillance at travel hubs a critical early warning tool.^2,3^ Wastewater-based epidemiology has been widely recognized as a valuable tool for monitoring pathogens of public health importance during the global SARS-CoV-2 pandemic^4–6^ and has been used to monitor respiratory viruses,^7^ zoonotic diseases,^8,9^ and enteroviruses.^10^ Wastewater sampling from airplanes is minimally invasive and non-disruptive to airport operations^11^ and allows for pathogen detection without engaging travelers. Modeling studies suggest that aviation wastewater surveillance, particularly on long-haul flights at select airports, can be an efficient and effective form of public health surveillance,^12^ and optimizing a strategic network of sentinel airports may optimize early warning detection of outbreaks.^13,14^

During the COVID-19 pandemic, several countries operationalized aviation wastewater pathogen surveillance to identify new and emerging SARS-CoV-2 variants and track trends in global spread.^15–21^ In the United States, the Centers for Disease Control and Prevention (CDC) has operated a Traveler-based Genomic Surveillance (TGS) program since 2021.^3^ The TGS program collects aviation wastewater samples and nasal swab samples from volunteer travelers arriving at international airports as part of an effort to track importation of emerging pathogens and variants. While nasal samples provide more granular information, aviation wastewater surveillance allows for pathogen detection without direct engagement of travelers.^11^

Similarly, in the United Kingdom, the UK Health Security Agency’s (UKHSA) Environmental Monitoring for Health Protection (EMHP) program led SARS-CoV-2 wastewater surveillance during the pandemic, including a three-week pilot in 2022 at three UK airports. This pilot, supported by the UK Home Office and academic partners, evaluated the use of wastewater from multiple collection points - including airplanes, terminal sewers, vacuum trucks, and wet wells - and demonstrated both the utility and complexity of pathogen surveillance in transport settings.^22–24^

Global aviation wastewater surveillance networks for early detection of pathogens have been previously suggested as a tool for pandemic preparedness and global biosecurity.^25^ Here, we present the results of an extended proof-of-concept pilot exploring feasibility of transatlantic airplane wastewater monitoring at international airports in the United States and the United Kingdom. The objective of this study was to assess the feasibility and analytical interoperability for early signal detection of public health threats using airplane wastewater monitoring at two international ports of entry. We highlight both the successes and limitations of implementing bilateral aviation wastewater surveillance in partnership with the airline industry.

## Results

### Polymerase chain reaction (PCR) detection

During 27 March 2024 – 16 January 2025, we collected 308 samples from flights originating in the United Kingdom and arriving in the United States. During 22 July 2024 – 15 January 2025, we collected 116 samples from flights originating in the United States and arriving in the United Kingdom (Figure 1). Of the 424 total airplane wastewater samples, 317 (74.8%) tested positive for at least one respiratory pathogen (influenza A virus, influenza B virus, SARS-CoV-2 or respiratory syncytial virus [RSV]) and 303 (71.5%) were positive for at least one enteric pathogen (norovirus GI, norovirus GII and adenovirus) (Figure 2).

**Figure 1.**
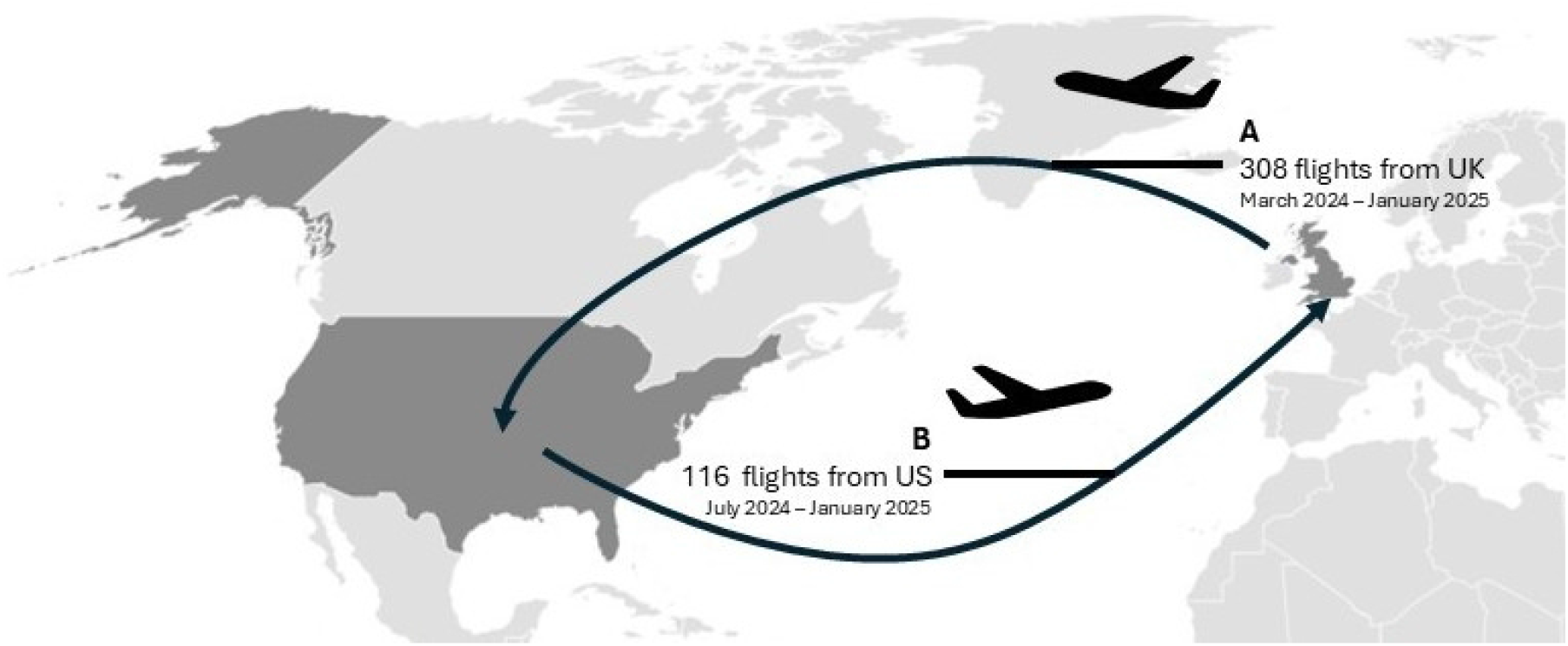
Overview of a bi-lateral partnership for aircraft wastewater surveillance, United Kingdom (UK) and United States (US), March 2024 -January 2025. 424 samples collected from multiple airlines along a common route were tested for seven pathogens, and sequenced for SARS-CoV-2.

**Figure 2.**
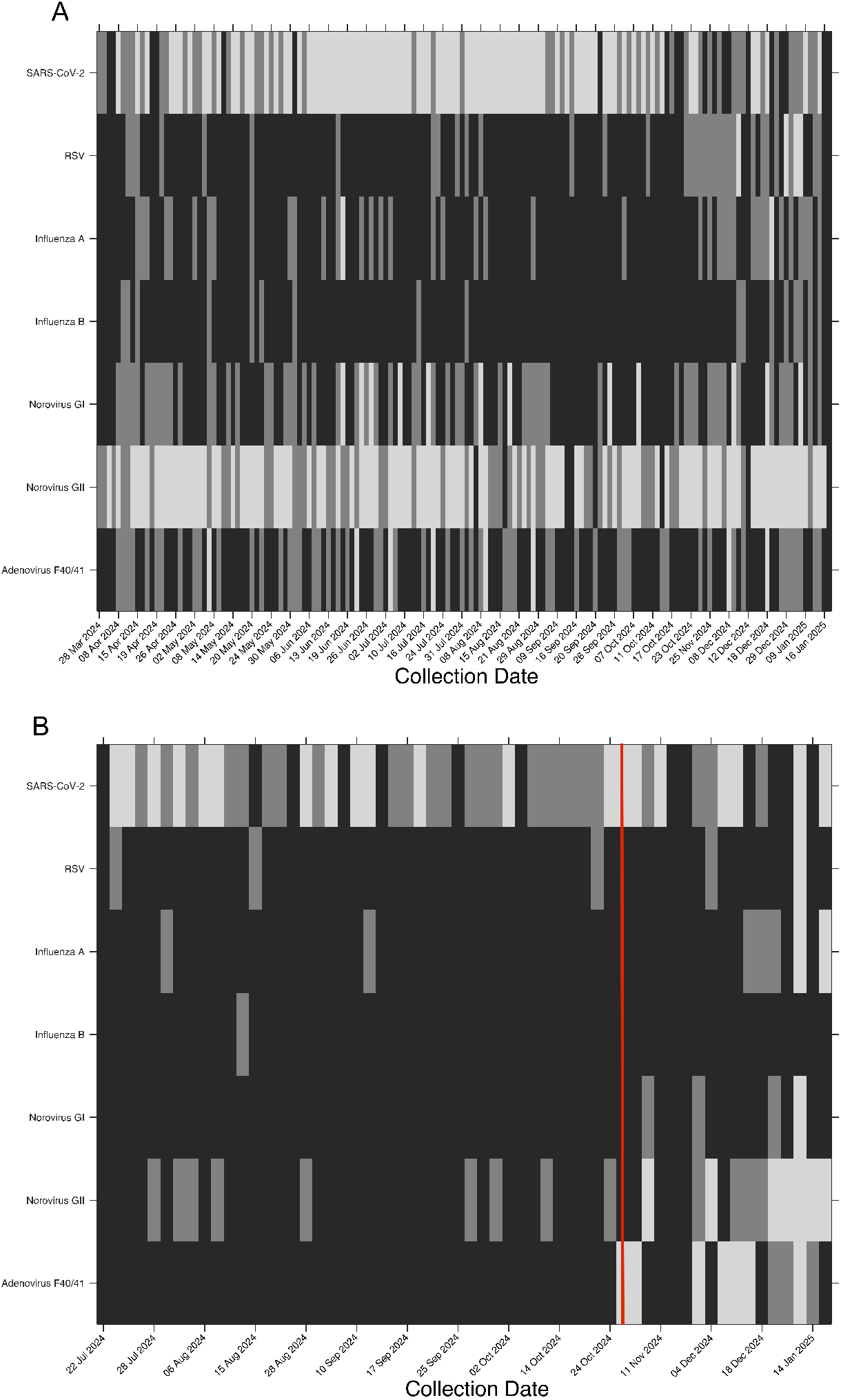
Heatmaps indicating detections of the seven targeted pathogens in airplane wastewater. per collection date, two flights per day per country, i.e., United Kingdom and United States, July 2024 - January 2025 (black = no detections, dark Gray = detection on one flight, light gray = detections on both flights); UK → US (A) and US → UK (B) flights. The red line indicates a change in laboratory for samples collected on US-origin flights, which occurred from 26 October 2024 onward. RSV = respiratory syncytial virus.

Norovirus GII returned the highest proportion of detections in UK-origin flights (79.9%), with a corresponding 23.3% detected from US-origin flights, followed by SARS-CoV-2 (72.4% UK, 56.0% US), norovirus GI (26.9% UK, 14.7% US origin), and adenovirus F40/41 (22.4% UK, 14.7% US) (Table 1). Influenza A virus positives were more frequently detected from flights from both origins towards the end of 2024, with UK origin samples detecting the virus marginally earlier (starting late November 2024) than US origin (starting the first half of December 2024), consistent with Northern Hemisphere seasonality (Figure 3).

**Table 1.** Digital polymerase chain reaction (dPCR) pathogen positivity of airplane wastewater samples (%) [total positives / total flights]. For samples collected from UK-origin flights and analysed by dPCR in the United States, results are presented with a standard threshold (T_S_) and after applying a three partition threshold (T_3P_). The change in dPCR positivity (%) indicates the percent reduction in positivity upon evaluating the standard threshold to the three positive partition threshold for each dPCR target. RSV = respiratory syncytial virus.

| Target | [T<sub>S</sub>] | [T<sub>3P</sub>] | % Change in Positivity |
| --- | --- | --- | --- |
| SARS-CoV-2 | 72.4% [223 / 308] | 47.4% [146 / 308] | -35% |
| RSV | 14.0% [43 / 308] | 5.2% [16 / 308] | -63% |
| Influenza A virus | 14.0% [43 / 308] | 4.5% [14 / 308] | -67% |
| Influenza B virus | 5.5% [17 / 308] | 1.9% [6 / 308] | -65% |
| Norovirus GI | 26.9% [83 / 308] | 8.8% [27 / 308] | -67% |
| Norovirus GII | 79.9% [246 / 308] | 56.8% [175 / 308] | -29% |
| Adenovirus F40/41 | 22.4% [69 / 308] | 11.7% [36 / 308] | -48% |

**Figure 3.**
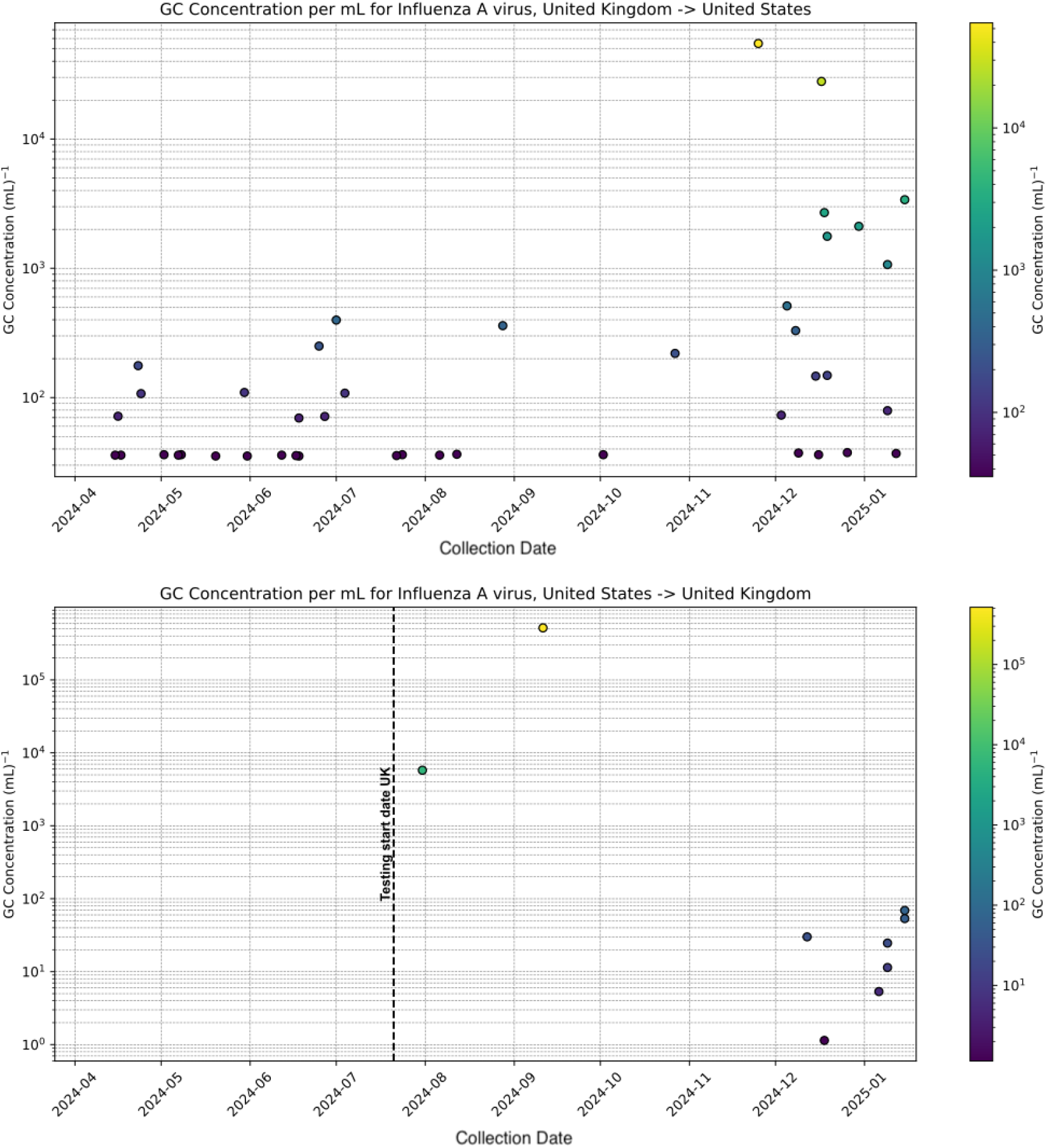
Scatter plots for gene copy (GC) concentrations of influenza A virus observed in airplane wastewater samples from UK-origin flights (top) and US-origin flights (bottom), April 2024 - January 2025. Note that samples from UK-origin flights were quantified using digital polymerase chain reaction (dPCR) and US-origin flights by quantitative PCR (qPCR) by two different laboratories (See Methods).

### Digital PCR threshold analysis

To evaluate the effect of detection thresholds, SARS-CoV-2 digital PCR (dPCR) results were classified under two criteria: a standard threshold (> 1 positive partition) and a more stringent threshold (>3 positive partitions). The whole genome sequencing for each sample was further evaluated and characterized by level of coverage and depth: ≥ 25%, ≥ 50%, and ≥ 70% coverage with a minimum of 10X depth.

Among samples designated as positive by the standard but negative by the 3-partition threshold, 80.5% (62/77) yielded SARS-CoV-2 genomes (with > 25% coverage with a minimum 10x read) demonstrating true positives despite lower partition counts (Table 2). No SARS-CoV-2 sequences were recovered from the 64 samples deemed negative by both thresholds supporting the specificity of detection. Applying a 3-partition cutoff across all dPCR targets would have significantly reduced measured positivity (Table 1).

**Table 2.** SARS-CoV-2 digital polymerase chain reaction (dPCR) results vs whole genome sequencing (WGS) results of airplane wastewater samples(%) [samples meeting WGS coverage and depth metric / total samples]. dPCR results were sorted into three categories: Positive under both thresholding approaches (standard (T_S_) and three positive partition (T_3P_)), positive with standard thresholding and negative with three positive partition thresholding, and negative with both thresholding approaches. For each dPCR category, WGS performance for those samples was evaluated using three criteria: ≥25%, ≥50%, and ≥70% coverage with a minimum of 10X depth.

| dPCR result |  | # of results* | Whole genome sequencing |  |  |
| --- | --- | --- | --- | --- | --- |
| [T<sub>S</sub>] | [T<sub>3P</sub>] |  | ≥ 25% coverage over 10X | ≥ 50% coverage over 10X | ≥ 70% coverage over 10X |
| Positive | Positive | 146 | 85.6% [125 / 146] | 84.2% [123 / 146] | 80.8% [118 / 146] |
| Positive | Negative | 77 | 80.5% [62 / 77] | 55.8% [43 / 77] | 37.7% [29 / 77] |
| Negative | Negative | 84 | 0% [0 / 64] | 0% [0 / 64] | 0% [0 / 64] |

*\*Table includes dPCR results for 307 samples, and sequencing results for 287 of the 308 samples from UK-origin flights. 20 samples were not sequenced and 1 sample was not tested via either dPCR or sequencing.*

### Genomic sequencing

SARS-CoV-2 lineage frequency showed parallel patterns across both directions of travel with the majority of BA.2.86 and its descendants (JN.1, KP.*, LB.*) (Figure 4). For both US-origin and UK-origin flights, KP.3.1.1* (47.8%; 35.0%, respectively) were the most common lineages captured. Other lineages of note included KP.2* (13.5% UK-origin, 10.0% US-origin) and KP.3* (19.6% UK=origin, 10.0% US-origin) lineages.

**Figure 4.**
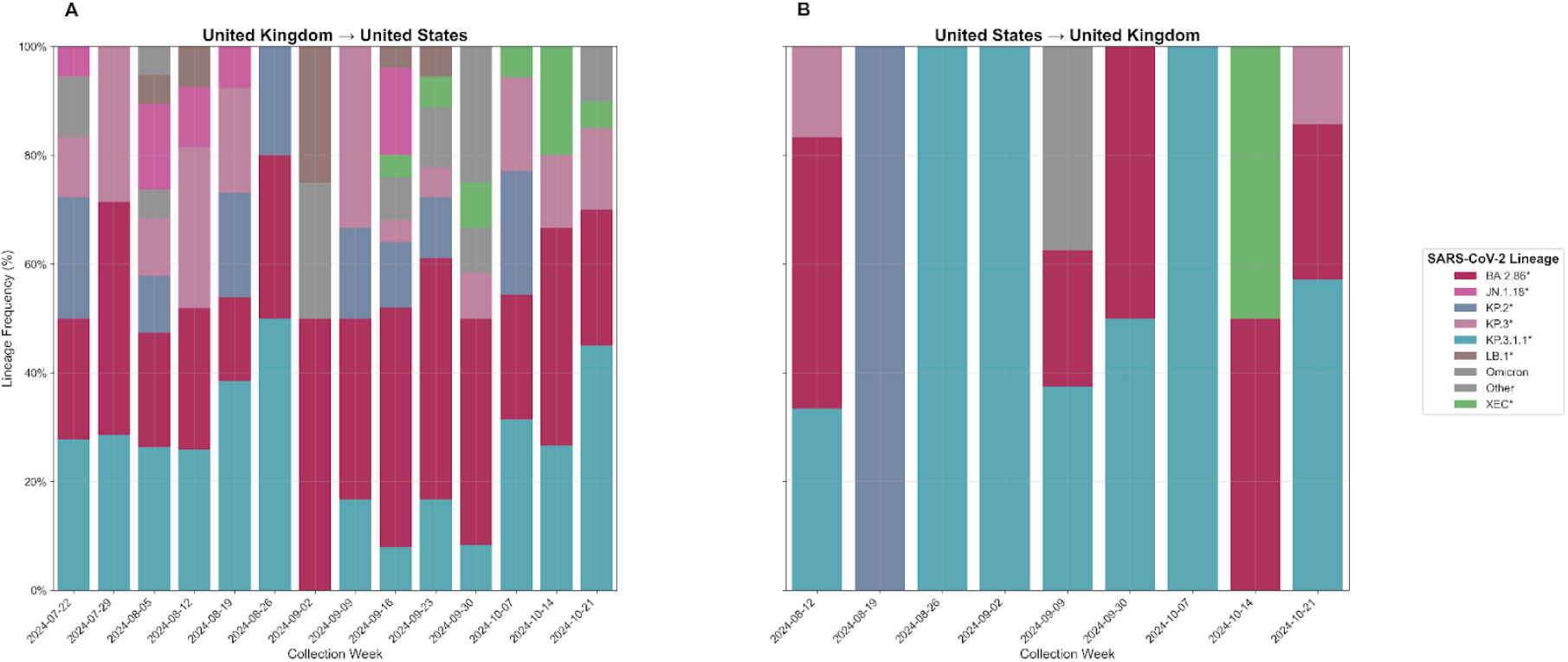
SARS-CoV-2 lineage frequency over time by country from airplane wastewater. Stacked bar charts showing the relative frequency of major SARS-CoV-2 lineages in samples from flights originating in the United Kingdom (A) and United States (B) from 23 July through 25 October 2024. Only samples with ≥70% reference coverage and assigned lineages are included (US-origin (63) and UK-origin (16)). Frequencies are calculated as percentages of total samples per collection week. Note change in sequencing process from Exeter Lab to UKHSA Lab. No sequences were obtained from US-origin flights after 21 October 2024.

Flights from the United Kingdom exhibited a greater lineage diversity, including JN.1.18* (5.6%) and LB.1* (4.7%), which were absent or rare in samples from US-origin flights.

### Detection of XEC

Notably, XEC* was detected 4 weeks earlier in samples from UK-origin flights, first appearing on 17 September, 2024, compared to US-origin flights on 14 October, 2024 (Figure 5). XEC defining mutations (S:F59S and A559T) were detected beginning in September 2024. Although the first US clinical case was reported earlier on 14 July 2024 (Virginia; GISAID: EPI_ISL_19310626) these airplane wastewater and subsequent community wastewater data suggest a lag in US community transmission, with the National Wastewater Surveillance System (NWSS) reporting >5% XEC only by 12 October 2024.^26^

**Figure 5.**
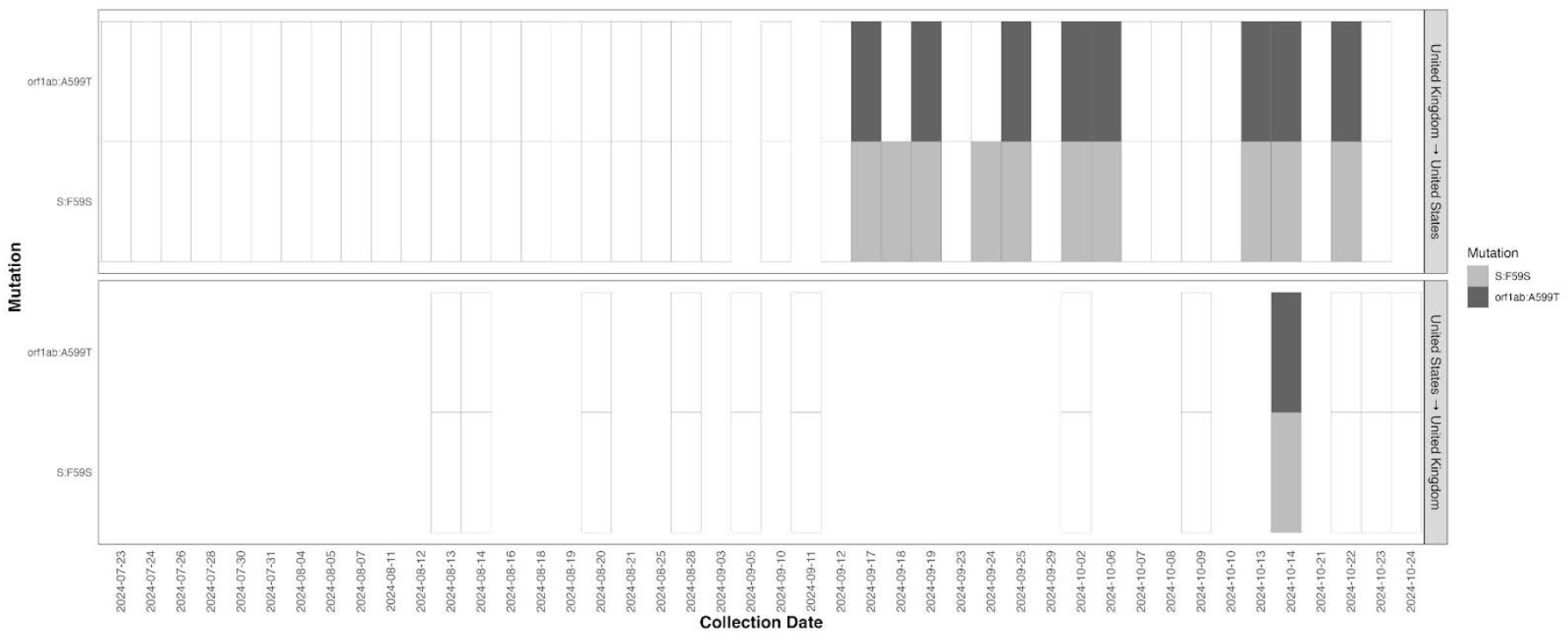
Testing of airplane wastewater from flights from the United Kingdom detected signals of SARS-CoV-2 variant XEC before flights from the United States. Sequencing results for UK-origin and US-origin samples from collection dates 23 July to 24 October 2024 (n=79)

## Discussion

This binational pilot demonstrated that airplane wastewater can be systemically collected and analyzed to advance global pathogen surveillance while maintaining passenger and airline privacy. Through a coordinated US–UK approach, we showed that bilateral data sharing can support early detection of pathogens and enables detection of trends at both origin and destination airports.

Our findings expand the role of near-source wastewater-based epidemiology complimenting surveillance in strategic locations such as educational institutions,^27,28^ correctional facilities,^29–31^ and healthcare settings.^32,33^ Aviation wastewater, which is rich in human waste and free of exogenous contributions, e.g. agricultural or industrial inputs, provides high-concentration signals and therefore can be a powerful early warning tool for pathogens of public health importance.^34^

Many wastewater surveillance programs use varied positivity thresholds depending on technical limitations and surveillance goals.^35,36^ Our finding that >25% genome coverage was achievable in samples with fewer than three positive dPCR partitions indicates that applying a more stringent threshold of three partitions may underestimate true positivity in aviation wastewater samples and delay detection of emerging variants or pathogens. Since 34.5% (77/223) of positive airplane wastewater samples fell below the three positive partition threshold, maintaining a lower threshold may maximize early warning potential when monitoring traveling populations.

The earlier detection of SARS-CoV-2 XEC in UK origin flights four weeks ahead of US origin flights and the recovery of diverse Omicron lineages (e.g. JN.1.18, LB.1) highlight our ability to utilize airplane wastewater samples to capture global lineage movement in near real time. Although XEC’s first clinical US case was documented in July 2024, airplane wastewater and subsequent community wastewater data indicate that significant US transmission lagged until mid-October underscoring the unique temporal sensitivity of aviation wastewater sampling. This is also supported by reports that XEC emerged in Europe and spread to the US later.^37,38^

This project illustrates that aviation industry partners are willing to collaborate on biosecurity and public health preparedness efforts when privacy and operational concerns are addressed. Aviation wastewater surveillance as part of national and global biosecurity and preparedness programs can improve early pathogen detection that can lead to rapid mitigation steps that help avoid costly border closures and travel bans such as those that impacted aviation industry during the COVID-19 pandemic.^39^ Key enablers for this program included anonymized collection protocols, coordination among public health agencies, transportation authorities, border health officials, and aviation industry partners; and clear agreements on data handling—elements that can guide similar programs elsewhere.

Several factors constrain interpretation of our findings. First, different PCR platforms were used: dPCR was utilized for samples from UK-origin flights tested in the United States and quantitative (qPCR) for samples from US-origin flights tested in the United Kingdom (until 25 October 2024) limiting direct quantitative comparison. Difference in assay sensitivity (dPCR has greater analytical sensitivity than qPCR in wastewater samples),^35^ may also account for the lower norovirus GII levels found on US-origin flights, especially as the US was experiencing an increase in norovirus outbreaks in late 2024.^40^ Second, sample size and temporal coverage differed (63 samples over 14 weeks from UK origin flights versus 16 samples over 9 weeks from US origin flights) which may explain the differences in SARS-CoV-2 lineage diversity. We could not fully assess seasonal patterns as the project lasted just under a year (March 2024 through January 2025). Finally, airplane wastewater measures viral shedding from an aggregation of passengers using airplane lavatories rather than prevalence. Therefore airplane wastewater surveillance provides an index of potential disease burden, with higher concentrations indicating a higher likelihood of multiple passenger pathogen carriage, while no detection (zero) indicating a high, but non-absolute, likelihood of pathogen absence.

This demonstration project suggests that expanding aviation wastewater surveillance by inclusion of additional international nodes, ensuring harmonization of assays and methods, and fostering multilateral data sharing could enhance global early warning detection of emerging pathogens. Recent multilateral exercises (e.g. European Commission’s GLOWACON Network^41^) confirm the feasibility of supranational wastewater sentinel systems.^25,42^ Scaling up such networks, integrated with genomic sequencing and rapid data exchange, could help detect and slow the spread of pathogens that could cause the next global pandemic.

## Methods

### Study design and site selection

Airplane wastewater samples were collected from flights traveling between an international airport in the United Kingdom and Dulles International Airport (IAD) in the United States over several months. Two major airlines operating a common transatlantic route participated in the study. Pathogen testing was conducted via polymerase chain reaction (PCR), and positive samples for SARS-CoV-2 were further analyzed using whole-genome sequencing.

To address industry privacy concerns and avoid stigmatizing specific airline operators, samples were deidentified at the point of collection. The US CDC, UKHSA and UK Department for Transport (DfT) developed safeguards to protect airline identities (e.g., no tail numbers, flight numbers, times of arrival or airline names were collected) while preserving critical public health data (e.g., country of origin, date of collection, pathogens detected, and sequencing results). This project was not intended as an evaluation of airline or airport hygiene practices, but instead focused on detection of infectious pathogens that might be circulating among travelers. This approach enabled the collection of meaningful public health data while minimizing the commercial or reputational risk to aviation partners.

### Sample collection and coordination

Wastewater (approximately 0.25 gal [1 L]) was collected from the lavatory waste tanks of the airplane during post arrival servicing. A vacuum device was attached to the airplane’s lavatory service panel port and connected to the lavatory service truck hose.

In the United States, collections began earlier due to existing airside access agreements under the CDC’s Traveler-based Genomic Surveillance (TGS) Program and continued one day beyond the UK collection window due to logistical constraints. Flights from the United Kingdom to the United States were sampled 27 March 2024 – 16 January 2025 at the US airport. Flights from the United States to the United Kingdom were sampled between 22 July 2024 – 15 January 2025 at the UK airport.

### Laboratory methods

Samples from UK-origin flights arriving in the United States were concentrated using Ceres Nanotrap^®^ Microbiome Particles and extracted using the Applied Biosystems™ MagMAX™ Microbiome Ultra Nucleic Acid Isolation Kit. Extracts were tested using the QIAGEN QIAcuity^®^ dPCR platform using QIAGEN QIAcuity^®^ OneStep Advanced Probe Kit on custom panels for SARS-CoV-2, adenovirus F, RSV, influenza A and B viruses, norovirus GI, and norovirus GII. SARS-CoV-2 whole genome sequencing following ARTICv5.3.2 protocol was performed using the Illumina^®^ NovaSeq™ X platform (paired end100 bp; Illumina).

Samples from US-origin flights were collected in the United Kingdom in two phases: phase 1 (22 July 2024 – 25 October 2024), when samples were processed using Ceres Nanotrap^®^ Microbiome Particles and extracted using the Applied Biosystems™ MagMAX™ Microbiome Ultra Nucleic Acid Isolation Kit. Extracts were tested using the Techne™ PrimePro 48 Real-Time PCR System with the Promega GoTaq^®^ Enviro RT-qPCR Kit for SARS-CoV-2, targeting the same panel of viruses listed above; phase 2 (26 October 2024 – 21 January 2025), when all samples were extracted using the Zymo Quick-DNA/RNA Water Kit, and RT-qPCR was performed using TaqManTM Fast Virus 1-Step Master Mix and TaqManTM Microbe Detection Assays on a QuantStudioTM 7 (ThermoFisher Scientific) for the panel of viruses listed above. Pepper Mild Mottle Virus (PMMoV) was used as a control and was detected using GoTaq® Enviro PMMoV Quant Kit (Promega). RNA was prepared for NGS library preparation using sample-independent single-primer amplification (SISPA). Libraries were prepared using the Illumina DNA Prep, (M) Tagmentation kit. Metagenomic sequencing of both DNA and RNA was performed on a NextSeq™ 2000 platform (Illumina).

### Bioinformatics and sequence analysis

Raw sequencing data were analyzed using a pipeline built in Nextflow (v22.04.5). Initial sequencing reads were demultiplexed (BCL2fastq; Illumina), reads filtered to those that pass Q30 ≥ 75% (fastqc v0.11.9), adaptors were trimmed (fastp v0.23.2), and human reads were removed (Kraken2 v2.1.2). Quality trimmed and filtered reads were aligned to the SARS-CoV-2 reference genome, MN908947.3, using bowtie2 (v2.4.4). Primers were removed (iVar v1.3.1), alignments were sorted and indexed (samtools v1.15.1), and duplicate reads were marked for high quality alignment (picard v2.27.4 + samtools v1.15.1). Variant calling and annotation was performed (freebayes v1.3.6 SnpEff v5.0, SnpSift v4.3), and a consensus genome was generated based on quality >100, depth >10, and alternative observations/depth >0.5 (bcftools v1.15.1 INFO/AO / INFO/DP > 0.5 & QUAL > 100 & INFO/DP > 10). Samples were deconvoluted for determining SARS-CoV-2 lineages and frequency esimates using Freyja. Consensus sequences were also generated and lineages were identified using Pangolin (v4.3.1). Tool parameter settings were used as default except for fastp (–cut_front –cut_tail –trim_poly_x –cut_mean_quality 30 –qualified_quality_phred 30 –unqualified_percent_limit 40 –length_required 50), freebayes (–ploidy 1 –min-alternate-fraction 0.1, –min-coverage 10), and bcftools (INFO/AO / INFO/DP > 0.5 & QUAL > 100 & INFO/DP > 10).

For qPCR, samples were considered positive if any amplification was observed. For dPCR, samples were considered positive if there was at least one positive partition. a quantitative comparison was made with samples having three or more positive partitions to assess threshold of detection. All samples that tested positive for SARS-CoV-2 by qPCR or dPCR were submitted for whole genome sequencing, regardless of Ct value or viral concentration, in accordance with our objective of evaluating the feasibility of aviation wastewater surveillance for early variant detection.

## Acknowledgments

Program participants; participating airlines and airports; the CDC Port Health Protection Branch and UKHSA Border Health team. We thank all Ginkgo Operations, Sequencing, and Epidemiology team members who contributed to this work, with special acknowledgment to past and present Program Managers: Claire Altieri, Ben H. Rome, Amy Schierhorn, Tim Lyden, and Robert C. Morfino. We acknowledge 20/30 Laboratories, UK (Mett Smart and Farzana Sultana) who conducted phase 1 of the UK experimental work.

## Author contributions statement

M.J.W., C.R.F., I.R., E.P., V.M., A.P.R., D.G. conceived the experiment(s), M.J.W., C.R.F., I.R., E.P., V.M., A.P.R., D.G., and the UKHSA Laboratory Team conducted the experiment(s), M.J.W., C.R.F., I.R., E.P., V.M., A.P.R., D.G., S.R., B.S. analysed the results. All authors reviewed the manuscript.

## Additional information

### Ethics and oversight

This activity was reviewed by CDC, deemed not research, and was conducted consistent with applicable federal law and CDC policy (see e.g., 45 C.F.R. part 46; 21 C.F.R. part 56; 42 U.S.C. §241(d), 5 U.S.C. §552a, 44 U.S.C. §3501 et seq.). The activity was also reviewed by the UKHSA Research & Public Health Practice Ethics and Governance Group, which deemed that there was no risk of harm to participants and no disclosure requirement due to the absence of personal information collected.

### Disclaimer

The findings and conclusions in this report are those of the authors and do not necessarily represent the official position of the Centers for Disease Control and Prevention, the UK Health Security Agency, or Department for Transport.

### Funding

This project has been funded by the Centers for Disease Control and Prevention (75D30122C14933) and the UK Health Security Agency.

### Competing interests

A.P.R., D.G., S.R., and B.S. were employed by Ginkgo Bioworks at the time of the study and received Ginkgo Bioworks stocks. M.J.W., I.R., E.P., V.M., and C.R.F. declare no competing interests.

### Data Availability

The datasets generated during and/or analysed during the current study are available in the following repositories:

- https://doi.org/10.5281/zenodo.18432347, and
- https://www.ncbi.nlm.nih.gov/bioproject/989177.

## Notes

### Funding Statement

This project has been funded by the Centers for Disease Control and Prevention (75D30122C14933). The UK Health Security Funded laboratory analysis of US-origin wastewater samples from the 26th October, 2024.

### Summary of Updates

Corrected the UKHSA Laboratory Team names to remove duplicates. Added “the UK Health Security Agency, or Department for Transport.” to the Disclaimer statement.

